# Characterizing multivariate regional hubs for schizophrenia classification, sex differences, and brain age estimation using explainable AI

**DOI:** 10.1101/2025.02.28.25323105

**Authors:** Yuzheng Nie, Taslim Murad, Hui-Yuan Miao, Puskar Bhattarai, Deepa S. Thakuri, Ganesh B. Chand

**Affiliations:** Department of Radiology, Mallinckrodt Institute of Radiology, Washington University School of Medicine, St. Louis, MO, USA; Institute for Informatics, Data Science and Biostatistics, Washington University School of Medicine, St. Louis, MO, USA; University of Missouri, School of Medicine, Columbia, MO, USA; Imaging Core, Knight Alzheimer Disease Research Center, Washington University School of Medicine, St. Louis, MO, USA; Institute of Clinical and Translational Sciences, Washington University School of Medicine, St. Louis, MO, USA; NeuroGenomics and Informatics Center, Washington University School of Medicine, St. Louis, MO, USA

**Author notes:** Correspondence: Dr. Ganesh B Chand and Dr. Hui-Yuan Miao. Co-first authors (Yuzheng Nie, Taslim Murad, Hui-Yuan Miao). Senior author (Ganesh B. Chand). **Credit authorship contribution statement** YN: Conceptualization, Methodology, Software, Formal analysis, Visualization, Data curation, Writing – original draft, Writing - review & editing; TM: Methodology, Software, Formal analysis, Writing - review & editing; HYM: Methodology, Writing – original draft, Writing - review & editing; PB: Methodology, Software, Writing – original draft, writing - review & editing; DST: Conceptualization, Data curation, Writing – original draft, writing - review & editing; GBC: Conceptualization, Methodology, Software, Formal analysis, Visualization, Data curation, Writing – original draft, Writing - review & editing; Supervision, Funding acquisition.

**Keywords:** Machine learning, Deep learning, Schizophrenia classification, Brain age prediction, Sex difference, and Shapley additive explanations

## Abstract

**Purpose:** To investigate multivariate regional patterns for schizophrenia (SZ) classification, sex differences, and brain age by utilizing structural MRI, demographics, and explainable artificial intelligence (AI).

**Methods:** Various AI models were employed, and the outperforming model was identified for SZ classification, sex differences, and brain age predictions. For the SZ and sex classification tasks, support vector classifier (SVC), k-nearest neighbor (KNN), and deep learning neural network (DL) models were compared. In the case of regression-based brain age prediction, Lasso regression (LR), Ridge regression (RR), support vector regression (SVR), and DL models were compared. For each regression or classification task, the optimal model was further integrated with the Shapley additive explanations (SHAP) and the significant multivariate brain regional patterns were identified.

**Results:** Our results demonstrated that the DL model outperformed other models in SZ classification, sex differences, and brain age predictions. We then integrated outperforming DL model with SHAP, and this integrated DL-SHAP was used to identify the individualized multivariate regional patterns associated with each prediction. Using DL-SHAP approach, we found that individuals with SZ had anatomical changes particularly in left pallidum, left posterior insula, left hippocampus, and left putamen regions, and such changes associated with SZ were different between female and male patients. Finally, we further applied DL-SHAP method to brain age prediction and suggested important brain regions related to aging in health controls (HC) and SZ processes.

**Conclusion:** This study systematically utilized predictive modeling and novel explainable AI approaches and identified the complex multivariate brain regions involved with SZ classification, sex differences, and brain aging and built a deeper understanding of neurobiological mechanisms involved in the disease, offering new insights to future SZ diagnosis and treatments and laying the foundation of the development of precision medicine.

## 1. Introduction

Schizophrenia (SZ) is widely regarded as one of the most debilitating health conditions affecting humanity [1-6], with lifetime prevalence chance of about 1% [7, 8], and is known to affect 1 in 300 individuals [9]. Males and females are affected by the disease differently [10-12]. SZ causes severe behavioral dysfunctions like hallucinations, delusions, and cognitive impairments in individuals, and it can accelerate brain aging processes by introducing brain alterations [13-22]. In contrast to the significant personal and socioeconomic burden cause by SZ, it remains challenging to reach a comprehensive understanding [23]. Despite extensive research and clinical efforts, effective personalized treatment options for SZ are still lacking. Therefore, building deeper understanding of SZ mechanisms, especially important multivariate brain regions associated with SZ, is crucial for improving diagnosis, developing more effective treatment strategies, and promoting precision medicine efforts.

Integrating magnetic resonance imaging (MRI) of the brain with machine learning/artificial intelligence (ML/AI) has dominated exploratory research lately [9, 15, 24-32] for SZ classification and age predictions. Existing studies have primarily focused on predictive modeling, but the underlying neurobiological regional patterns associated with such predictions are poorly understood yet. Prior research examining which brain regions are closely related to SZ has primarily used a univariate approach, neglecting the multivariate associations between regions. Since different brain regions work in a collaborative way and various regions have been implicated with SZ mechanisms [33-38], it is critical to systematically investigate the associations between the brain regions and SZ mechanisms in a multivariate way, thereby uncovering the complex neurobiological mechanisms of this disorder.

This study sought to investigate the underlying multivariate regional patterns associated with neurobiological mechanisms, focusing on SZ vs. control classification, as well as male vs. female classification and brain age predictions in SZ using a large sample of MRI and demographic data. For this, we built various AI models for these predictions and integrated the outperforming model with the feature importance method to explain/interpret the predictions and identify the corresponding multivariate regional patterns. Since individuals with SZ exhibit brain regional changes [24, 39-44], we hypothesize that our explainable AI approach can predict SZ vs. control classification and identify the multivariate regional contributors to such prediction. Since male vs. female disparity has been suggested in SZ [12, 45, 46], we hypothesize that our explainable AI-based modeling can characterize the neurostructural correlates in a multivariate fashion. Since some studies suggest the effect of age with SZ [14, 47], we further test this using our explainable AI approach.

## 2. Materials and Methods

### 2.1 Dataset

The dataset (N = 368, Age: 18-66 years old, **Table 1**) was obtained from Schizconnect [48, 49], including data from following projects: Center for Biomedical Research Excellence (COBRE) [50], Neuromorphometry by Computer Algorithm Chicago (NMorphCH) [51], and function Biomedical Informatics Research Network (fBIRN) PhaseII 0010 [52]. Out of 368 subjects, 165 (42 females) subjects belong to SZ group, and 203 (79 females) are healthy controls (HC). We utilized the brain T1-weighted MR images and extracted brain volumes corresponding to 145 anatomical regions of interest (ROIs) using the MUlti-atlas region Segmentation utilizing Ensembles of registration algorithms and parameters and locally optimal atlas selection (MUSE) [53]. These brain volumes were further corrected for the site and covariate effects using harmonization technique [4, 54]. Using this harmonization approach, the sex and age effects were corrected for SZ classification, age effects were corrected for sex classification, and sex effects were corrected for brain age prediction. These harmonized brain regional volumes corresponding to 145 ROIs were utilized as inputs to the ML/DL models. The 145 ROIs include a wide range of tissue types, including gray matter, white matter, cerebrospinal fluid/ventricles, and brain stem.

**Table 1:**
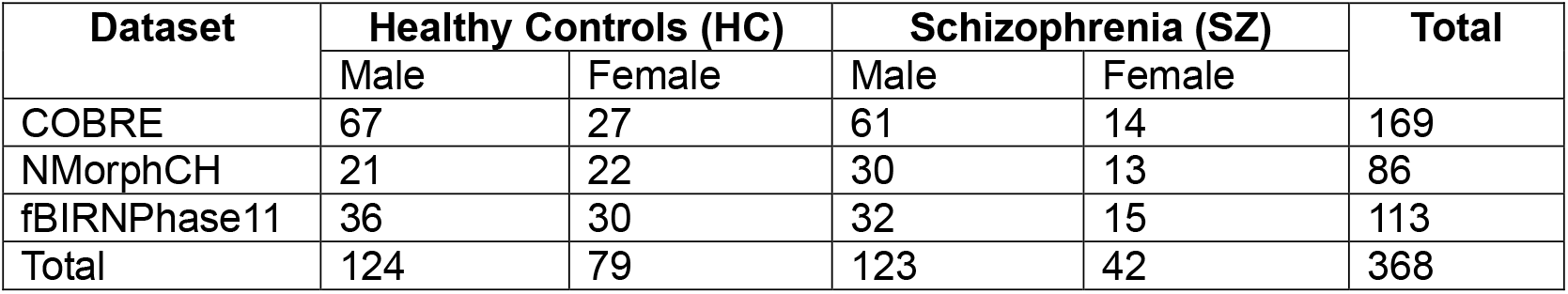
Demographic distribution of participants.

### 2.2 SZ-related classification and regression tasks and associated AI models

We evaluated 3 SZ-related tasks, including two classification tasks and one regression task. The classification tasks included classifying SZ vs. HC and classifying male vs. female sex within SZ group and HC group. We employed traditional ML models, including K-nearest neighbor (KNN) [37, 38] and support vector classifier (SVC) [39, 40], along with a deep learning neural networks (DL) model for classification tasks. The regression task estimated the brain age of SZ subjects., We employed traditional ML models, including Lasso regression (LR) [42, 43], Ridge regression (RR) [44, 45], and support vector regression (SVR) [55], as well as DL model for the regression task.

The hyperparameters of traditional ML models were tuned with a grid search procedure, and the optimal hyperparameters were selected for each model and each classification or regression task. For KNN, the optimal number of neighbors was chosen from a range of 1-100 with an increment of 2. For SVC, the gamma value was chosen from -12 to -2 with an increment of 1. In addition, the Gaussian/Radial basis function (RBF) kernel was employed, and we evaluated C values ranging from 0.2 to 2.6 with an increment of 0.2. For LR and RR, the optimal value for alpha parameter was selected from the bounds 0.15-0.4 and 50-250, respectively. For SVR, an RBF kernel was employed and the optimal values of gamma, epsilon, and c-values hyperparameters were selected from a range of -12 to -2 with an increment of 1, -7 to 3 with an increment of 1, and -5 to 4 with an increment of 1, respectively.

We evaluated a DL model consisting of five dense hidden layers with decreasing number of units in order of 200, 160, 120, 80, and 40. Each dense layer was followed by a rectified linear unit (ReLU) activation function, a batch normalization layer, and a dropout layer with 0.1 dropout rate to stabilize the model and accelerate the training process [56, 57]. The final output layer comprises a single-unit dense layer accompanied by a linear activation function for regression task and a sigmoid activation function for classification task. This architecture was chosen as it yielded the optimal predictions for both regression and classification tasks.

To ensure the robustness of our models, a 10-fold cross-validation (CV) strategy was implemented, where data were split into train and test sets. The train set was further divided into train and validation sets, where validation set was 10% of the original train set, to monitor the training loss during the CV-based iterative training. CV is critically acclaimed for enhancing model generalization, mitigating overfitting, and providing a more robust estimation of the model’s performance on unseen data [58]. The outcomes reported here are from the partitioned test set, which were not used in training the models. The traditional ML models were implemented in Python using Python library Scikit-learn [59] and trained with the default settings. To train the DL model, binary cross entropy (BCE) loss function was employed for classification and mean absolute error (MAE) loss function for regression tasks. After hyperparameter tuning, we selected 500 training epoch, 64 batch size, 0.001 learning rate, and ADAM optimizer. The results for DL model were reported after running the model multiple times (i.e., five times here) and averaging the results over runs for the robustness purposes. For classification, a soft voting approach was used to get averaged outputs. The DL model was implemented using Python libraries TensorFlow [60] and Keras [61].

Various AI models were compared for each classification or regression task and the outperforming model (i.e., DL model in our study here) was integrated with the Shapley additive explanation (SHAP) [62], referred to as DL-SHAP, to explain or interpret the model outputs, thereby identifying the significant multivariate regional contributions to each classification or regression task. SHAP approach was utilized in this study because it is a state-of-the-art strategy for the interpretability or explainability of complex models and it satisfies all three essential properties, particularly local accuracy, missingness, and consistency, that are critical for the interpretability or explainability and accuracy of complex AI models. SHAP comprises model-specific approximations, integrates the strengths of all other additive feature attribution methods, and outperforms other methods.

This integration concept was originally proposed to investigate multivariate associations between brain regions and cognition in the context of Alzheimer disease [63], and here we intended to apply it to schizophrenia domain. In DL-SHAP, the SHAP value for an input feature is computed by considering the contribution it makes to the final output. This contribution can be determined by observing the impact on final output when the feature is added and when it’s removed from the input. A higher absolute SHAP value indicates greater significance of the feature in obtaining the output. In our study, a higher absolute SHAP value associated with a ROI indicates a stronger association between that brain region and the related prediction. Note that the feature significance values with DL-SHAP across the models might vary as the contribution of the features on these models might change due to variations in architecture, weights, and the complexity of the model. Hence, the feature significance values with DL-SHAP are best used for understanding relative feature contributions within the same model, rather than for direct comparison across different models [62].

For assessing the classification model, we used micro averaged accuracy, precision, recall, specificity, and F1-score as evaluation metrics. Note that, reporting multiple evaluation metrics can provide deeper insights about the models’ performance. For regression tasks, we computed the spearman correlation between the actual and predicted brain age, and the related p-values to evaluate model performance. Cohen’s d effect sizes were computed from DL-SHAP values to examine group differences. Group-wise DL-SHAP regional features and DL-SHAP group differences in terms of Cohen’s d regional values were visualized using MRIcroGL [64]. Only ROIs with *p*-values less than 0.05, indicating statistical significance, were considered.

## 3. Results

For the SZ vs. HC classification task, we found that DL model outperformed (loss = 0.206, accuracy = 0.981, precision = 0.998, recall = 0.970, F1-Score = 0.979, Specificity = 0.990) all other traditional ML models in all evaluation metrics. The results of HC/SZ classification using various ML and DL models are listed in **Table 2**.

**Table 2:** SZ vs. HC classification. The test set performance of models. The optimal value for each evaluation metric is marked bold. KNN: K-nearest neighborhood; SVC: support vector classifier; DL: Deep learning neural networks.

| Model | Accuracy | Precision | Recall | F1-Score | Specificity | Loss |
| --- | --- | --- | --- | --- | --- | --- |
| KNN | 0.601 | 0.615 | 0.291 | 0.395 | 0.852 | 13.810 |
| SVC | 0.652 | 0.653 | 0.479 | 0.552 | 0.793 | 11.526 |
| DL | **0.981** | **0.988** | **0.970** | **0.979** | **0.990** | **0.206** |

We then characterized the multivariate relationships between regional volumetric measures and HC/SZ classification using DL-SHAP. We identified the important brain regions for HC and SZ groups and their classification task. In HC group, the top important regions were the left pallidum, left posterior insula, left hippocampus, fornix left, and left putamen, among others (**Figure 1a**). In SZ group the top important regions identified were the left pallidum, left posterior insula, left hippocampus, right pallidum, and left putamen, among others (**Figure 1b**). SHAP values at individual participant level associated with the top important regions were visualized in **Figure S1**, which demonstrated the relationships between the volumetric measurements of individual ROI and SZ. For example, a larger pallidum is associated with a higher likelihood of being SZ patients and a smaller insula and hippocampus are associated with a higher likelihood of being SZ patients. We then computed Cohen’s d between HC and SZ groups using DL-SHAP regional features and found few regions (**Figure 1c**) that were not key hubs in each group.

**Figure 1:**
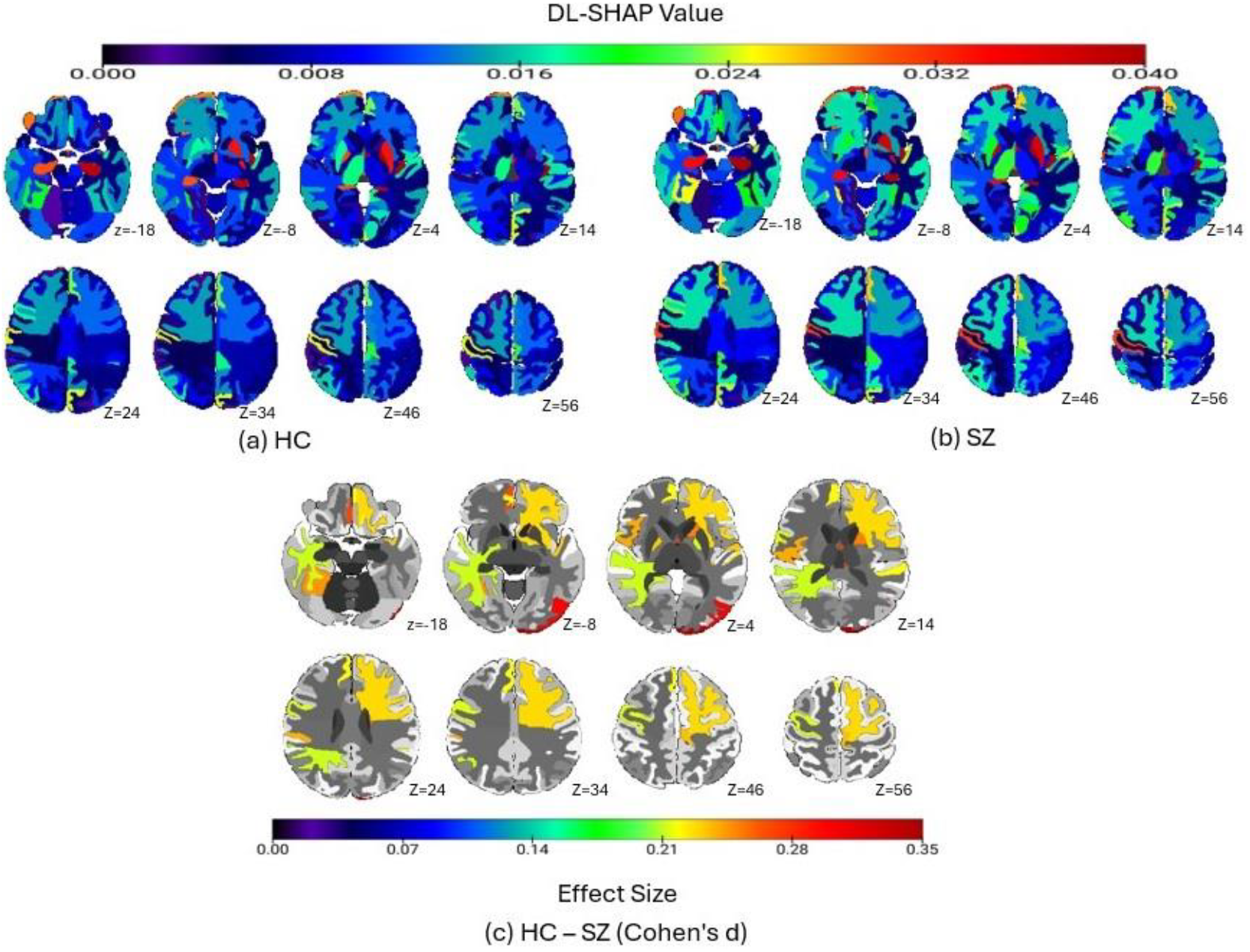
SZ vs. HC diagnosis classification via DL-SHAP. (a) shows significant brain regions in HC group. (b) illustrates significant brain regions in SZ. (c) portrays the Cohen’s d analysis comparing HC and SZ brains. The color bar represents the absolute value of effect size.

Sex classification was performed within HC group and SZ group, respectively. Our results indicated that the DL model outperformed (SZ group: loss = 0.188, accuracy = 0.994, precision = 0.992, recall = 1, F1-Score = 0.996, specificity = 0.976; HC group: loss = 0.162, accuracy = 0.995, precision = 1.000, recall = 0.992, F1-Score = 0.996, specificity = 1.000) all other traditional ML models corresponding to all evaluation metrics. The results of sex classification using various ML and DL models are listed in **Table 3** and **Table 4**. We further investigated the multivariate relationships between regional volumetric measures and sex classification. The top important regions identified for male in HC group were the left temporal pole, right posterior insula, left ventral DC, left hippocampus, and left entorhinal area (**Figure 2a**); for male in SZ group were the left posterior cingulate gyrus, left cuneus, right posterior orbital gyrus, left inferior temporal gyrus, and right inferior temporal gyrus (**Figure 2b**); for female in HC group were the left temporal pole, left ventral DC, left entorhinal area, right posterior insula, and right occipital lobe white matter (**Figure 2c**); and for female in SZ group were left posterior cingulate gyrus, corpus callosum, left inferior temporal gyrus, 4th ventricle, and right posterior orbital gyrus (**Figure 2d**). We compared sex differences within each of HC and SZ groups with the Cohen’s d analysis (HC: **Figure 2e**; SZ: **Figure 2f**).

**Table 3:** Sex-wise classification within SZ group. The test set performance of classification models. The optimal value for each evaluation is marked bold.

| Model | Accuracy | Precision | Recall | F1-Score | Specificity | Loss |
| --- | --- | --- | --- | --- | --- | --- |
| KNN | 0.800 | 0.836 | 0.911 | 0.871 | 0.476 | 6.869 |
| SVC | 0.824 | 0.851 | 0.927 | 0.887 | 0.524 | 2.390 |
| DL | **0.994** | **0.992** | **1.000** | **0.996** | **0.976** | **0.188** |

**Table 4:** Sex-wise classification within HC group. The test set performance of models. The optimal value for each evaluation metric is marked bold.

| Model | Accuracy | Precision | Recall | F1-Score | Specificity | Loss |
| --- | --- | --- | --- | --- | --- | --- |
| KNN | 0.778 | 0.801 | 0.847 | 0.823 | 0.671 | 7.639 |
| SVC | 0.749 | 0.774 | 0.831 | 0.801 | 0.620 | 4.218 |
| DL | **0.995** | **1.000** | **0.992** | **0.996** | **1.000** | **0.162** |

**Figure 2:**
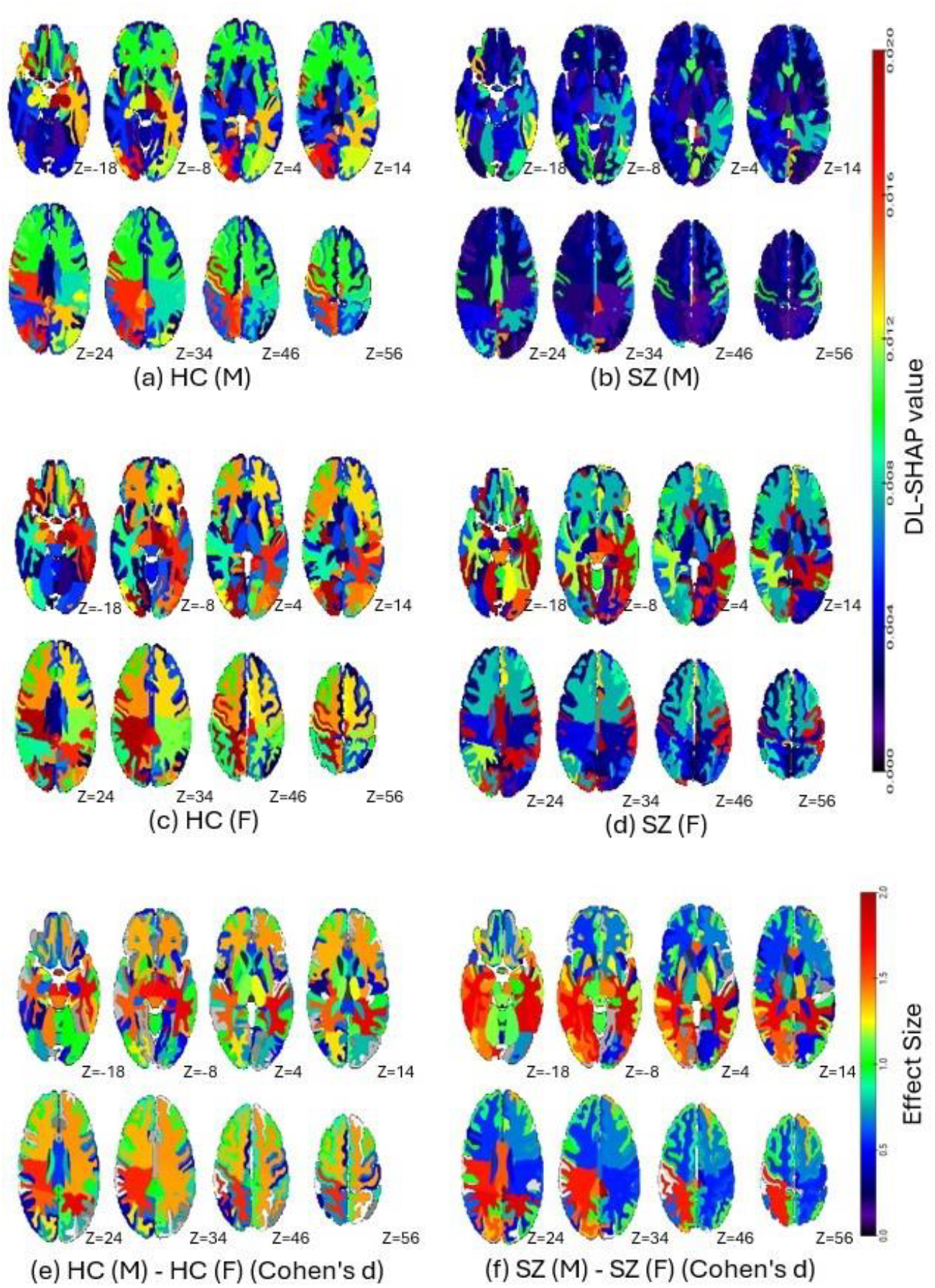
Sex-wise classification within HC and SZ groups via DL-SHAP model. (a) shows significant brain regions in males within HC group for sex classification. (b) illustrates significant brain regions in males for sex-classification within SZ group. (c) demonstrates significant brain regions in females for sex classification within HC group. (d) represents significant brain regions in females within SZ. (e) shows the Cohen’s d analysis (absolute value) comparing male and female brains in HC group. (f) shows the Cohen’s d analysis (absolute value) comparing male and female brains in SZ group.

Finally, we found that DL model outperformed all other traditional ML models in all evaluation metrics with a loss of 3.96 and a correlation of 0.92 (p = 7.9E-150) between the actual and predicted brain age (**Figure 3a**). On contrast, the traditional ML models yielded loss around 7 and correlation lower than 0.7. The results of brain age prediction using various ML and DL models are listed in **Table 5**. The multivariate relationships between the regional measures and brain age in SZ were also investigated using DL-SHAP. The top important regions identified for HC group were the right superior frontal gyrus, left putamen, left supplementary motor cortex, right entorhinal area, and right subcallosal area (**Figure 3c)**, and the top important regions identified for SZ group were the right superior frontal gyrus, left putamen, left pallidum, right subcallosal area, and right entorhinal area (**Figure 3d**). **Figure 3b** showed the Cohen’s d effect size differences between HC and SZ groups using DL-SHAP regional features.

**Table 5:** Brain age prediction. The test set performance of regression models in terms of MAE training loss, Spearman’s correlation between actual and prediction brain age, and *p*-values. The optimal value for each evaluation metric is marked bold. LR: lasso regression; RR: ridge regression; SVR support vector regression; DL: deep learning neural networks.

| Model | MAE Loss | Spearman's Correlation | P-value |
| --- | --- | --- | --- |
| LR | 6.91 | 0.67 | 5.84E-49 |
| RR | 6.97 | 0.68 | 2.66E-50 |
| SVR | 7.53 | 0.60 | 1.03E-38 |
| DL | **3.97** | **0.92** | **7.9E-150** |

**Figure 3:**
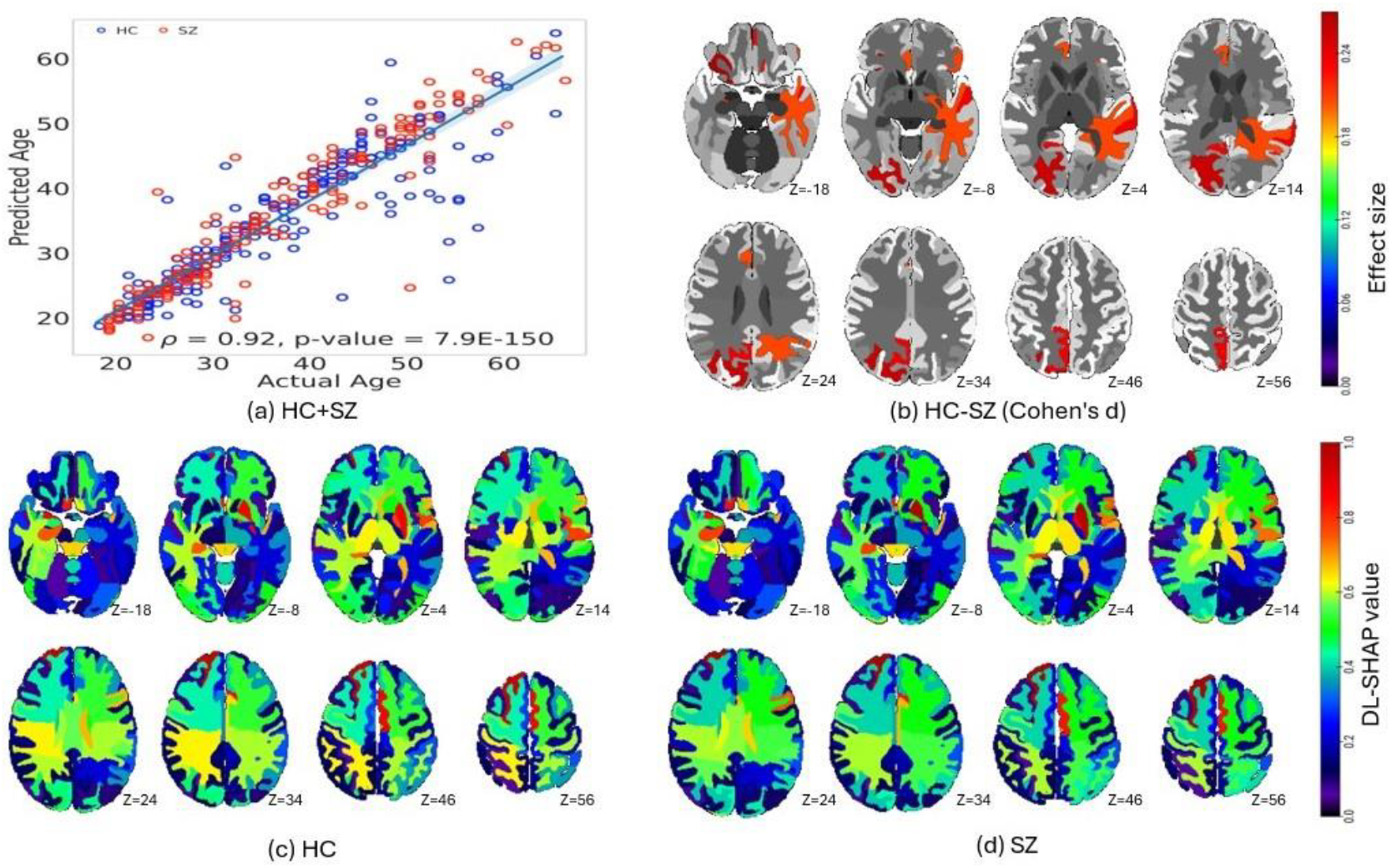
Brain age prediction via DL-SHAP model. (a) displays the correlation between actual and predicted brain age. (b) shows the Cohen’s d analysis (absolute value) comparing HC and SZ brains. (c) represents the significant brain regions for HC group. (d) represents the significant brain regions for SZ group.

## 4. Discussion

This study systematically investigated the hierarchy of multivariate brain regions associated with SZ mechanisms, particularly HC/SZ classification, sex classification, and brain age prediction. We found that the DL model outperformed the rest of ML models in all classification- and regression-based predictions, and this finding was broadly consistent with previous studies [56, 57, 65-68] suggesting superior performance of DL model for classification and brain age predictions in various disorders including SZ. The DL model provided a reliable model with decent performance for our purpose of investigating the multivariate relationships between regional brain biomarkers and different aspects of SZ. We then integrated the outperforming DL with SHAP, collectively referred to as DL-SHAP. The integrated DL-SHAP explainable AI approach uncovered that the individuals with SZ had anatomical changes, particularly in left pallidum, left posterior insula, left hippocampus, and left putamen regions, and such brain alterations associated with SZ showed a different pattern in female and male patients.

We found important brain regions in terms of DL-SHAP regional features for HC and SZ classification task, suggesting those regions undergo structural changes with the development of SZ. The identified key brain regions largely overlap with existing literature [69-74]. For example, subcortical structural abnormalities in SZ were previously studied and the left pallidum and left putamen volumetric increases were reported in SZ [71]. Our results showed larger pallidum and putamen are associated with a higher likelihood of being an SZ patient. Moreover, hippocampus and insula abnormalities were also reported in SZ [72, 73]. Our results showed smaller insula and hippocampus are associated with a higher likelihood of being an SZ patient. The broad agreement between our findings and those existing literature supports the validity of our results, but we innovatively identified the hierarchy of multivariate regions by considering their interactions using our novel DL-SHAP method for SZ vs. HC classification.

Previous studies have showed that the susceptibility of males and females to SZ is different [12, 45, 46]. We extended those findings by examining sex differences in a fine-grained and multivariate regional manner. A general consensus in the field is that SZ is associated with reduced frontal and temporal volumes in male than female [46]. We identified that left inferior temporal gyrus and right posterior orbital gyrus are important for sex classification in both male and female patients. In addition, similar to previous studies [75], we also identified left posterior cingulate gyrus as an important region for sex classification. Finally, ventricles and corpus callosum were shown to be different between male and female SZ patients [46]. In our study, they were identified as important regions for female SZ patients, but not as important for male SZ patients. While previous studies demonstrated some brain regions impacted differently in male and female by SZ, our study went one step further by showing both the importance of the same brain regions to sex classification is different for male and female, and different brain regions contribute to sex classification in male and female SZ patients. These results and existing literature collectively suggested the complex interplay between sex and SZ, and such complex neurobiological regional patterns could be innovatively identified using our explainable AI approach in a multivariate way.

Finally, for brain age prediction, our findings about significant brain regions also overlapped with the existing literature. For instance, prior reports [47, 76] suggest that SZ has a strong impact on aging process in right superior frontal gyrus and putamen. While an increase in the volume of entorhinal area was shown to be associated with SZ [77], it is new that our study explicitly demonstrated its aging process to be closely related with SZ. These findings and existing literature taken together suggested the impact of aging in HC and SZ processes.

One limitation of our study is that there are different subtypes in SZ [24, 78-82] but we could not investigate the multivariate relationships between different brain regions and different subtypes of due to the limitation of sample size. Future studies may consider employing a larger dataset, grouping participants by their SZ subtypes and investigate the multivariate relationships between regional brain biomarkers and different SZ subtypes.

## 5. Conclusion

In conclusion, we utilized a range of ML/DL models to systematically investigate SZ neurobiological mechanisms, focusing on SZ classification, sex differences, and brain age using MRI and demographic data. The DL model outperformed other models in all classification and regression tasks. Our integrated novel DL-SHAP method further provided valuable insights into the dominant multivariate regional brain hubs associated with SZ diagnosis, sex-based differences, and brain age prediction. These findings collectively contribute to a deeper understanding of the underlying neurobiological mechanisms of SZ, offering new perspectives that could aid in diagnostic, prognostic, and therapeutic strategies. This study is expected to serve as the foundation when these explainable AI approaches are extended to other neuroimage modalities, like fMRI, PET, and EEG, among others for the multifactorial holistic understanding of SZ and other related disorders.

## Data Availability

All data produced are available online at:
Cobre data was downloaded from the COllaborative Informatics and Neuroimaging Suite Data Exchange tool (COINS; http://coins.mrn.org/dx)
The fBIRN data used for this study were downloaded from the Function BIRN Data Repository (http://fbirnbdr.birncommunity.org:8080/BDR/)
NMorhCH data used in preparation of this article were obtained from the Neuromorphometry by Computer Algorithm Chicago (NMorphCH) dataset (http://nunda.northwestern.edu/nunda/data/projects/NMorphCH).

http://fbirnbdr.birncommunity.org:8080/BDR/

http://nunda.northwestern.edu/nunda/data/projects/NMorphCH

## List of abbreviations

(SZ): Schizophrenia
(AI): Artificial intelligence
(SVC): Support vector classifier
(KNN): K-nearest neighbor
(DL): Deep learning neural network
(LR): Lasso regression
(RR): Ridge regression
(SVR): Support vector regression
(SHAP): Shapley additive explanations
(HC): Healthy controls
(ML): Machine learning
(MRI): Magnetic resonance imaging
(MUSE): MUlti-atlas region Segmentation utilizing Ensembles of registration algorithms and parameters and locally optimal atlas selection
(ROIs): Regions of interest
(RBF): Radial basis function
(ReLU): Rectified linear unit
(CV): Cross-validation
(BCE): Binary cross entropy
(MAE): Mean absolute error.

## Acknowledgments

Schizconnect data collection and sharing for this project was funded by NIMH cooperative agreement 1U01 MH097435. Cobre data was downloaded from the COllaborative Informatics and Neuroimaging Suite Data Exchange tool (COINS; http://coins.mrn.org/dx) and data collection was performed at the Mind Research Network and funded by a Center of Biomedical Research Excellence (COBRE) grant 5P20RR021938/P20GM103472 from the NIH to Dr. Vince Calhoun. The fBIRN data used for this study were downloaded from the Function BIRN Data Repository (http://fbirnbdr.birncommunity.org:8080/BDR/), supported by grants to the Function BIRN (U24-RR021992) Testbed funded by the National Center for Research Resources at the National Institutes of Health, U.S.A. NMorhCH data used in preparation of this article were obtained from the Neuromorphometry by Computer Algorithm Chicago (NMorphCH) dataset (http://nunda.northwestern.edu/nunda/data/projects/NMorphCH). As such, the investigators within NMorphCH contributed to the design and implementation of NMorphCH and/or provided data but did not participate in analysis or writing of this report and data collection and sharing for this project was funded by NIMH grant R01 MH056584.

## References

1. McCutcheon, R.A., T.R. Marques, and O.D. Howes, Schizophrenia-An Overview. Jama Psychiatry, 2020. 77(2): p. 201–210.

2. Crespo-Facorro, B., et al., The burden of disease in early schizophrenia - a systematic literature review. Curr Med Res Opin, 2021. 37(1): p. 109–121.

3. Chand, G.B., et al., Differential Sphingosine-1-Phosphate Receptor-1 Protein Expression in the Dorsolateral Prefrontal Cortex Between Schizophrenia Type 1 and Type 2. Frontiers in Psychiatry, 2022. 13.

4. Chand, G.B., et al., Schizophrenia Imaging Signatures and Their Associations With Cognition, Psychopathology, and Genetics in the General Population. American Journal of Psychiatry, 2022. 179(9): p. 650–660.

5. Insel, T.R., Rethinking schizophrenia. Nature, 2010. 468(7321): p. 187–193.

6. Tandon, R., et al., The schizophrenia syndrome, circa 2024: What we know and how that informs its nature. Schizophrenia Research, 2024. 264: p. 1–28.

7. Zhu, C., et al., Temporal Dynamic Synchronous Functional Brain Network for Schizophrenia Classification and Lateralization Analysis. IEEE Trans Med Imaging, 2024. PP.

8. Adamu, M.J., et al., Unraveling the pathophysiology of schizophrenia: insights from structural magnetic resonance imaging studies. Frontiers in Psychiatry, 2023. 14.

9. Kanyal, A., et al., Multi-modal deep learning from imaging genomic data for schizophrenia classification. Frontiers in Psychiatry, 2024. 15.

10. Galderisi, S., et al., No gender differences in social outcome in patients suffering from schizophrenia. Eur Psychiatry, 2012. 27(6): p. 406–8.

11. Schultz, S.H., S.W. North, and C.G. Shields, Schizophrenia: a review. Am Fam Physician, 2007. 75(12): p. 1821–9.

12. Salehi, M.A., et al., Brain-based sex differences in schizophrenia: A systematic review of fMRI studies. Hum Brain Mapp, 2024. 45(5): p. e26664.

13. Koutsouleris, N., et al., Accelerated brain aging in schizophrenia and beyond: a neuroanatomical marker of psychiatric disorders. Schizophr Bull, 2014. 40(5): p. 1140–53.

14. Shahab, S., et al., Brain structure, cognition, and brain age in schizophrenia, bipolar disorder, and healthy controls. Neuropsychopharmacology, 2019. 44(5): p. 898–906.

15. Bashyam, V.M., et al., MRI signatures of brain age and disease over the lifespan based on a deep brain network and 14 468 individuals worldwide. Brain, 2020. 143: p. 2312–2324.

16. Shen, C.-L., et al., Progressive brain abnormalities in schizophrenia across different illness periods: a structural and functional MRI study. Schizophrenia, 2023. 9(1): p. 2.

17. DeLisi, L.E., et al., Understanding structural brain changes in schizophrenia. Dialogues Clin Neurosci, 2006. 8(1): p. 71–8.

18. Shenton, M.E., et al., A review of MRI findings in schizophrenia. Schizophrenia Research, 2001. 49(1): p. 1–52.

19. Cheng, Y., et al., Brain Network Localization of Gray Matter Atrophy and Neurocognitive and Social Cognitive Dysfunction in Schizophrenia. Biological Psychiatry, 2025. 97(2): p. 148–156.

20. Xu, R., et al., Brain structural damage networks at different stages of schizophrenia. Psychological Medicine, 2024: p. p1-11.

21. McCutcheon, R.A., R.S.E. Keefe, and P.K. McGuire, Cognitive impairment in schizophrenia: aetiology, pathophysiology, and treatment. Molecular Psychiatry, 2023. 28(5): p. 1902–1918.

22. Jiang, S., et al., Progressive trajectories of schizophrenia across symptoms, genes, and the brain. BMC Medicine, 2023. 21(1): p. 237.

23. Tandon, R., et al., The schizophrenia syndrome, circa 2024: What we know and how that informs its nature. Schizophrenia Research, 2024. 264: p. 1–28.

24. Chand, G.B., et al., Two distinct neuroanatomical subtypes of schizophrenia revealed using machine learning. Brain, 2020. 143(3): p. 1027–1038.

25. Chilla, G.S., et al., Machine learning classification of schizophrenia patients and healthy controls using diverse neuroanatomical markers and Ensemble methods. Sci Rep, 2022. 12(1): p. 2755.

26. Zhang, J., et al., Detecting schizophrenia with 3D structural brain MRI using deep learning. Sci Rep, 2023. 13(1): p. 14433.

27. Priyanka, G., et al., Early Diagnosis of Schizophrenia in Patients Using Deep Learning Techniques. 2024 10th International Conference on Advanced Computing and Communication Systems (ICACCS), 2024. 1: p. 809–815.

28. Lee, W.H., et al., Brain age prediction in schizophrenia: Does the choice of machine learning algorithm matter? Psychiatry Research-Neuroimaging, 2021. 310.

29. Kim, W.S., et al., Deep Learning-based Brain Age Prediction in Patients With Schizophrenia Spectrum Disorders. Schizophr Bull, 2024. 50(4): p. 804–814.

30. Haker, R., et al., Characterization of Brain Abnormalities in Lactational Neurodevelopmental Poly I:C Rat Model of Schizophrenia and Depression Using Machine-Learning and Quantitative MRI. J Magn Reson Imaging, 2024.

31. Di Camillo, F., et al., Magnetic resonance imaging–based machine learning classification of schizophrenia spectrum disorders: a meta-analysis. Psychiatry and Clinical Neurosciences, 2024. 78(12): p. 732–743.

32. Chatterjee, I. and B. Hilal, Investigating the association between symptoms and functional activity in brain regions in schizophrenia: A cross-sectional fmri-based neuroimaging study. Psychiatry Research: Neuroimaging, 2024. 344: p. 111870.

33. Chand, G.B., I. Hajjar, and D.Q. Qiu, Disrupted interactions among the hippocampal, dorsal attention, and central-executive networks in amnestic mild cognitive impairment. Human Brain Mapping, 2018. 39(12): p. 4987–4997.

34. Chand, G.B., D.S. Thakuri, and B. Soni, Salience network anatomical and molecular markers are linked with cognitive dysfunction in mild cognitive impairment. Journal of Neuroimaging, 2022. 32(4): p. 728–734.

35. Chand, G.B., et al., Interactions of the Salience Network and Its Subsystems with the Default-Mode and the Central-Executive Networks in Normal Aging and Mild Cognitive Impairment. Brain Connectivity, 2017. 7(7): p. 401–412.

36. Chand, G.B. and M. Dhamala, The salience network dynamics in perceptual decision-making. Neuroimage, 2016. 134: p. 85–93.

37. Chand, G.B. and M. Dhamala, Interactions between the anterior cingulate-insula network and the fronto-parietal network during perceptual decision-making. Neuroimage, 2017. 152: p. 381–389.

38. Thakuri, D.S., et al., Dysregulated Salience Network Control over Default-Mode and Central-Executive Networks in Schizophrenia Revealed Using Stochastic Dynamical Causal Modeling. Brain Connect, 2024. 14(1): p. 70–79.

39. Shenton, M.E., T.J. Whitford, and M. Kubicki, Structural neuroimaging in schizophrenia: from methods to insights to treatments. Dialogues Clin Neurosci, 2010. 12(3): p. 317–32.

40. Haijma, S.V., et al., Brain volumes in schizophrenia: a meta-analysis in over 18 000 subjects. Schizophr Bull, 2013. 39(5): p. 1129–38.

41. Clementz, B.A., et al., Identification of Distinct Psychosis Biotypes Using Brain-Based Biomarkers. Am J Psychiatry, 2016. 173(4): p. 373–84.

42. Gupta, C.N., et al., Patterns of Gray Matter Abnormalities in Schizophrenia Based on an International Mega-analysis. Schizophr Bull, 2015. 41(5): p. 1133–42.

43. Koutsouleris, N., et al., Structural correlates of psychopathological symptom dimensions in schizophrenia: a voxel-based morphometric study. Neuroimage, 2008. 39(4): p. 1600–12.

44. Rozycki, M., et al., Multisite Machine Learning Analysis Provides a Robust Structural Imaging Signature of Schizophrenia Detectable Across Diverse Patient Populations and Within Individuals. Schizophr Bull, 2018. 44(5): p. 1035–1044.

45. Li, X., W. Zhou, and Z. Yi, A glimpse of gender differences in schizophrenia. Gen Psychiatr, 2022. 35(4): p. e100823.

46. Mendrek, A. and A. Mancini-Marïe, Sex/gender differences in the brain and cognition in schizophrenia. Neuroscience & Biobehavioral Reviews, 2016. 67: p. 57–78.

47. Zhu, J.-D., et al., Investigating brain aging trajectory deviations in different brain regions of individuals with schizophrenia using multimodal magnetic resonance imaging and brain-age prediction: a multicenter study. Translational Psychiatry, 2023. 13(1): p. 82.

48. Wang, L., et al., SchizConnect: Mediating neuroimaging databases on schizophrenia and related disorders for large-scale integration. Neuroimage, 2016. 124(Pt B): p. 1155–1167.

49. Ambite, J.L., et al., SchizConnect: Virtual Data Integration in Neuroimaging. Data Integr Life Sci, 2015. 9162: p. 37–51.

50. Aine, C.J., et al., Multimodal Neuroimaging in Schizophrenia: Description and Dissemination. Neuroinformatics, 2017. 15(4): p. 343–364.

51. Neuromorphometry by Computer Algorithm Chicago (NMorphCH). http://nunda.northwestern.edu/nunda/data/projects/NMorphCH.

52. Keator, D.B., et al., The Function Biomedical Informatics Research Network Data Repository. Neuroimage, 2016. 124(Pt B): p. 1074–1079.

53. Doshi, J., et al., MUSE: MUlti-atlas region Segmentation utilizing Ensembles of registration algorithms and parameters, and locally optimal atlas selection. Neuroimage, 2016. 127: p. 186–195.

54. Fortin, J.P., et al., Harmonization of cortical thickness measurements across scanners and sites. Neuroimage, 2018. 167: p. 104–120.

55. Chand, G.B., et al., Estimating regional cerebral blood flow using resting-state functional MRI via machine learning. J Neurosci Methods, 2020. 331: p. 108528.

56. Lam, P., et al., Accurate brain age prediction using recurrent slice-based networks. bioRxiv, 2020.

57. Kuo, C.Y., et al., Improving Individual Brain Age Prediction Using an Ensemble Deep Learning Framework. Front Psychiatry, 2021. 12: p. 626677.

58. Kohavi, R., A study of cross-validation and bootstrap for accuracy estimation and model selection, in Proceedings of the 14th international joint conference on Artificial intelligence - Volume 2. 1995, Morgan Kaufmann Publishers Inc.: Montreal, Quebec, Canada. p. 1137–1143.

59. Pedregosa, F., et al., Scikit-learn: Machine Learning in Python. Journal of Machine Learning Research, 2011.

60. Abadi, M., et al. TensorFlow: A system for large-scale machine learning. in USENIX Symposium on Operating Systems Design and Implementation. 2016.

61. Gulli, A. and S. Pal, Deep learning with Keras. 2017: Packt Publishing Ltd.

62. Lundberg, S.M. and S.-I. Lee. A Unified Approach to Interpreting Model Predictions. in Neural Information Processing Systems. 2017.

63. Bhattarai, P., et al., Explainable AI-based Deep-SHAP for mapping the multivariate relationships between regional neuroimaging biomarkers and cognition. Eur J Radiol, 2024. 174: p. 111403.

64. MRIcroGL software. https://www.nitrc.org/projects/mricrogl.

65. Gong, W., et al., Optimising a Simple Fully Convolutional Network for Accurate Brain Age Prediction in the PAC 2019 Challenge. Front Psychiatry, 2021. 12: p. 627996.

66. Oh, J., et al., Identifying Schizophrenia Using Structural MRI With a Deep Learning Algorithm. Frontiers in Psychiatry, 2020. 11.

67. Ahmedt-Aristizabal, D., et al., Identification of Children at Risk of Schizophrenia via Deep Learning and EEG Responses. Ieee Journal of Biomedical and Health Informatics, 2021. 25(1): p. 69–76.

68. Srinivasagopalan, S., et al., A deep learning approach for diagnosing schizophrenic patients. Journal of Experimental & Theoretical Artificial Intelligence, 2019. 31(6): p. 803–816.

69. Ito, S., et al., Association between globus pallidus volume and positive symptoms in schizophrenia. Psychiatry Clin Neurosci, 2022. 76(11): p. 602–603.

70. Yasuda, Y., et al., Brain morphological and functional features in cognitive subgroups of schizophrenia. Psychiatry Clin Neurosci, 2020. 74(3): p. 191–203.

71. Tang, Y.L., et al., Striatum and globus pallidus structural abnormalities in schizophrenia: A retrospective study of the different stages of the disease. Progress in Neuro-Psychopharmacology & Biological Psychiatry, 2024. 133.

72. Wegrzyn, D., G. Juckel, and A. Faissner, Structural and Functional Deviations of the Hippocampus in Schizophrenia and Schizophrenia Animal Models. Int J Mol Sci, 2022. 23(10).

73. Sheffield, J.M., et al., Insula functional connectivity in schizophrenia. Schizophr Res, 2020. 220: p. 69–77.

74. Tohid, H., M. Faizan, and U. Faizan, Alterations of the occipital lobe in schizophrenia. Neurosciences (Riyadh), 2015. 20(3): p. 213–24.

75. Lang, X.-E., et al., Sex difference in association of symptoms and white matter deficits in first-episode and drug-naive schizophrenia. Translational Psychiatry, 2018. 8(1): p. 281.

76. Ballester, P.L., et al., Gray matter volume drives the brain age gap in schizophrenia: a SHAP study. Schizophrenia, 2023. 9(1): p. 3.

77. Baiano, M., et al., Decreased entorhinal cortex volumes in schizophrenia. Schizophr Res, 2008. 102(1-3): p. 171–80.

78. Wen, J., et al., Nine Neuroimaging-AI Endophenotypes Unravel Disease Heterogeneity and Partial Overlap across Four Brain Disorders: A Dimensional Neuroanatomical Representation. medRxiv, 2024.

79. du Plessis, S., et al., Two Neuroanatomical Signatures in Schizophrenia: Expression Strengths Over the First 2 Years of Treatment and Their Relationships to Neurodevelopmental Compromise and Antipsychotic Treatment. Schizophr Bull, 2023. 49(4): p. 1067–1077.

80. Dwyer, D.B., et al., Psychosis brain subtypes validated in first-episode cohorts and related to illness remission: results from the PHENOM consortium. Mol Psychiatry, 2023. 28(5): p. 2008–2017.

81. Hwang, G., et al., Assessment of Neuroanatomical Endophenotypes of Autism Spectrum Disorder and Association With Characteristics of Individuals With Schizophrenia and the General Population. JAMA Psychiatry, 2023. 80(5): p. 498–507.

82. Wen, J., et al., Multi-scale semi-supervised clustering of brain images: Deriving disease subtypes. Med Image Anal, 2022. 75: p. 102304.

