## Supplemental material for "Characterizing multivariate regional hubs for schizophrenia classification, sex differences, and brain age estimation using explainable AI"

### Supplementary

#### Feature importance of HC/SZ classification task

We identified top regions with a high impact on HC/SZ classification task. **Figure S1** shows the top 20 regions drives the classification results, decided by their mean absolute SHAP value across all participants. The most important regions are on the top of the plot. This result is highly consistent across all runs, and here we showed results based on the first run. In our analysis, left pallidum showed a high feature importance in HC/SZ classification. Higher volume of pallidum is associated with higher SHAP value, indicating higher volume of pallidum is associated with higher likelihood of being an SZ patient. The second important region is the left posterior insula, and opposite to pallidum, having a smaller insula is associated with a higher likelihood of having SZ. Those results are highly consistent with the existing literature [1, 2].

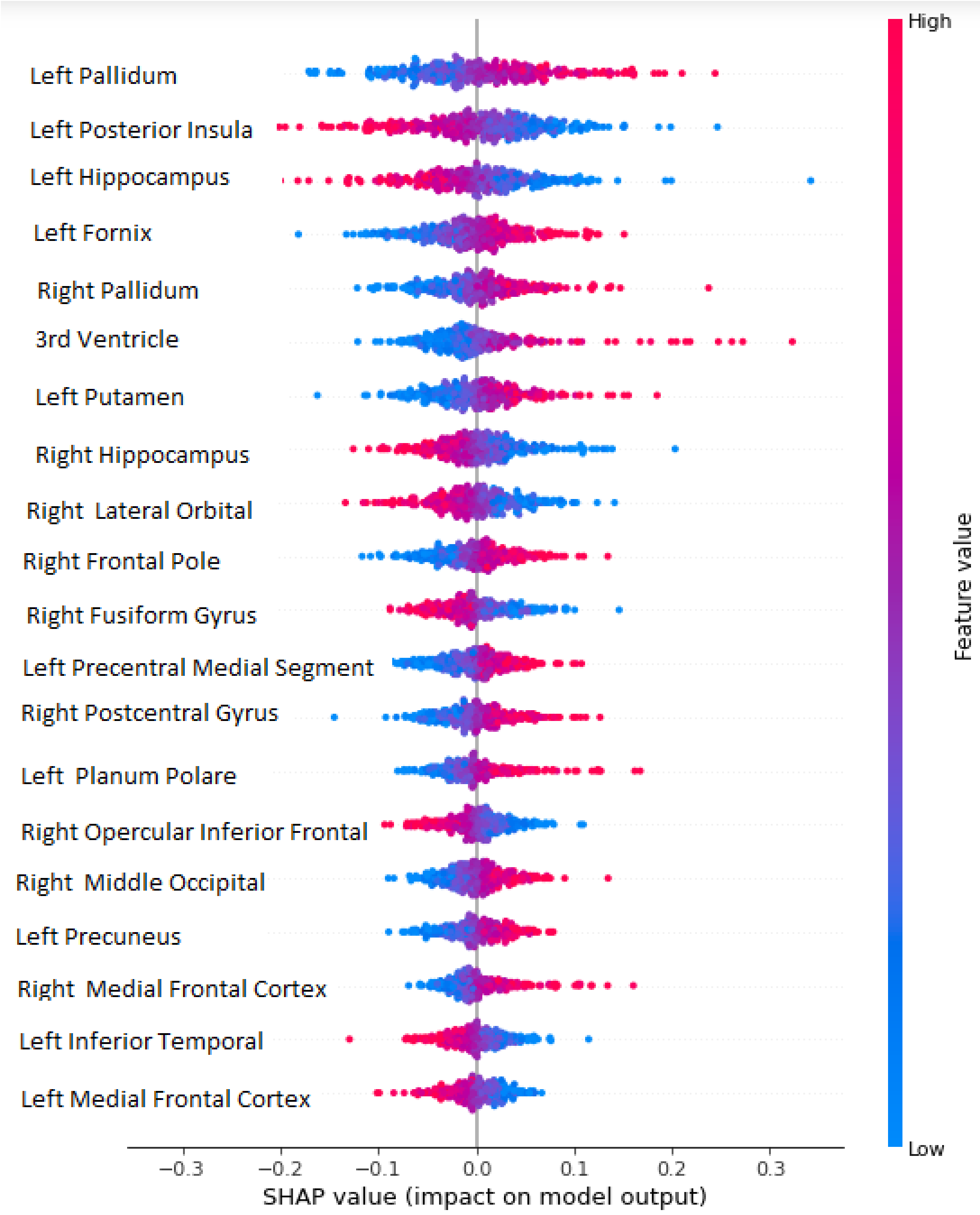


**Figure S1: SHAP value associated with different brain region in HC/SZ classification task**. Brain regions were ranked by their mean absolute SHAP values across all participants with a descending order such that the most important region is plotted at the top. Each dot represents an individual participant, and a higher SHAP value is associated with a higher likelihood of being an SZ patient. The color of dots is associated with the feature value, the volumetric measurements of brain regions, with blue being small regional volume and red being high regional volume.

### References

1. Sheffield, J.M., et al., *Insula functional connectivity in schizophrenia.* Schizophr Res, 2020. **220**: p. 69-77.
2. Tang, Y.L., et al., *Striatum and globus pallidus structural abnormalities in schizophrenia: A retrospective study of the different stages of the disease.* Progress in Neuro-Psychopharmacology & Biological Psychiatry, 2024. **133**.
